# Wastewater Surveillance of Oncogenic Viruses: A Baseline Assessment in Southeast Queensland, Australia

**DOI:** 10.64898/2026.09.02.26362014

**Authors:** Regina Keller, Metasebia Gebrewold, Wendy Smith, Rory Verhagen, Stuart L. Simpson, Catherine Hoar, Hannah Greenwald Healy, Warish Ahmed

## Abstract

Wastewater surveillance (WS) offers a non-invasive means of tracking population-level circulation of infectious agents, including viruses linked to cancer. This study provides the first Australian assessment of oncogenic viruses in municipal wastewater by screening 76 influent samples collected over four months from six wastewater treatment plants in Southeast Queensland, Australia. Ten gene targets representing seven oncogenic viruses— Epstein–Barr virus (EBV), hepatitis B virus (HBV), hepatitis C virus (HCV), human herpesvirus 8 (HHV-8), human papillomavirus 16 and 18 (HPV-16 and 18), human T-lymphotropic virus type 1 (HTLV-1), and Merkel cell polyomavirus (MCPyV)—were analysed using PCR-based methods. All viruses were detected in wastewater at least once, though with substantial variation in frequency. MCPyV was the most frequently detected virus, appearing in 97.3% of samples with concentrations ranging from 3.09-3.85 log₁₀ gene copies (GC)/50 mL, indicating widespread population exposure. HBV (26.3%) and EBV (15.8%) were detected intermittently across multiple catchments, while HPV-16/18, HHV-8, HTLV-1, and HCV were detected at the lowest frequencies (<8%). This study reports the first baseline dataset for oncogenic viruses in Australian wastewater. More broadly, positive detection of all targeted oncogenic viruses—including those associated with low-prevalence infections—in wastewater demonstrates the potential of WS to complement existing cancer surveillance systems in tracking community-level circulation of these infectious agents.

**Graphical abstract:** 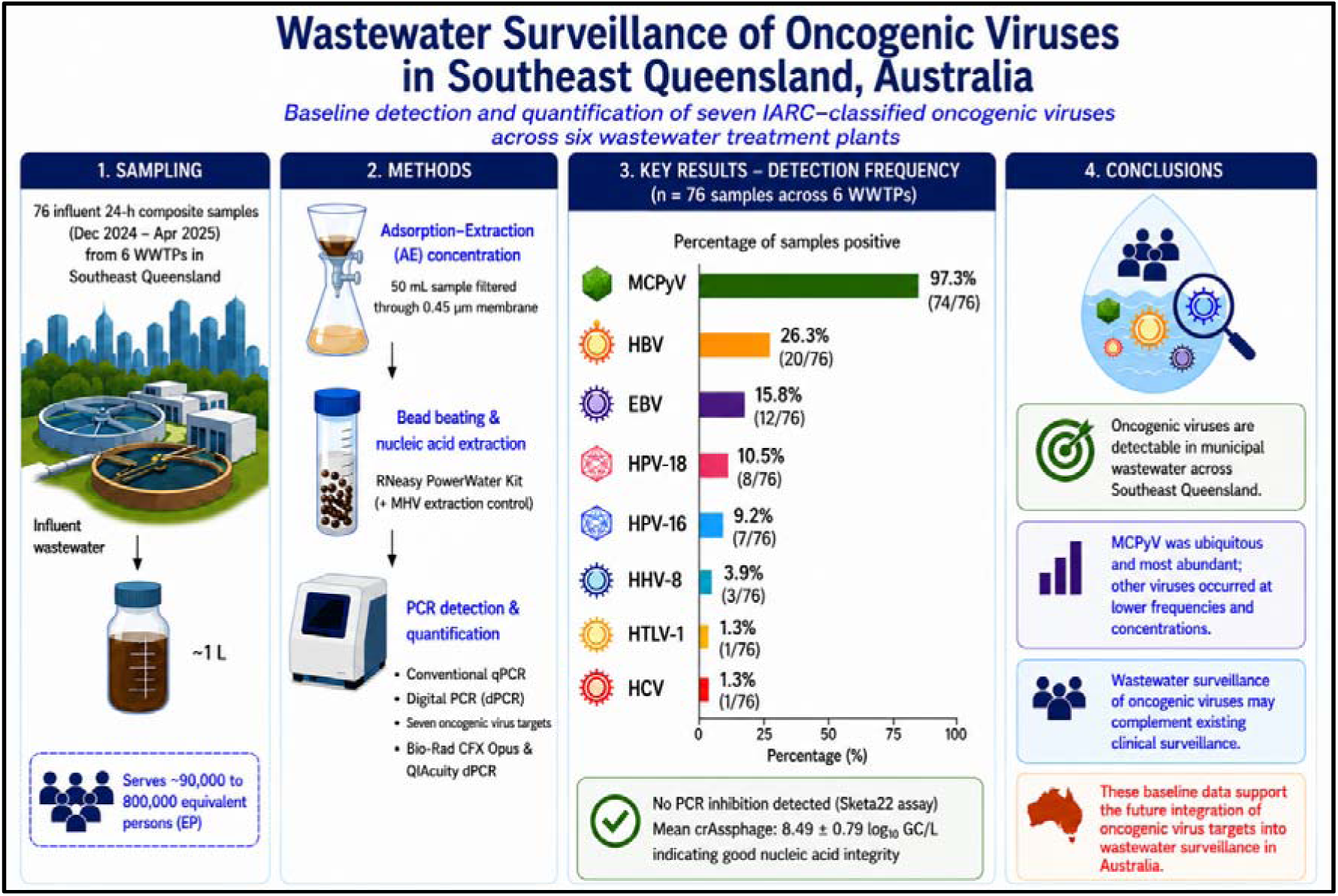

**Highlights:**

- Seven oncogenic viruses were screened in influent wastewater from six WWTPs.
- MCPyV was detected with the highest frequency, in 97.3% of samples.
- HBV and EBV were detected intermittently across multiple WWTP catchments.
- HPV, HHV-8, HTLV-1 and HCV were detected at the lowest frequencies.
- Wastewater surveillance shows promise for monitoring oncogenic viruses.

## 1. Introduction

Cancer is a leading cause of morbidity and mortality worldwide, with an estimated 20 million new cases reported in 2022 and projections rising to 35 million by 2050 (Bray et al., 2024). Approximately 15-20% of the global cancer burden is attributable to infectious agents (Zur Hausen, 2009), of which viruses account for an estimated 8-10% of all cancer cases (De Martel et al., 2020). Infection-related cancers remain an important public health challenge worldwide, , although their burden varies considerably between countries (Torre et al., 2026), depending on access to vaccination, screening and early diagnostic services.

Seven human oncogenic viruses (oncogene carriers) are classified as Group 1 carcinogens by the International Agency for Research on Cancer (IARC, 2012) (Table 1). These include human papillomavirus (HPV) (Ang et al., 2010), hepatitis B and C viruses (HBV and HCV) (El-Serag, 2011), Epstein–Barr virus (EBV) (Young and Rickinson, 2004), human T-cell lymphotropic virus type 1 (HTLV-1) (Chlichlia and Khazaie, 2010), and human herpesvirus 8 (HHV-8), also referred to as Kaposi sarcoma-associated herpesvirus (KSHV) (Mesri et al., 2010). Each of these oncoviruses are associated with specific malignancies which manifest through diverse mechanisms, including insertional mutagenesis, chronic inflammation, immune modulation, and disruption of host cell cycle regulation (Butel, 2000).

**Table 1.** Human oncogenic viruses included in this study and their characteristics relevant to wastewater surveillance.

| Virus | Genome type | Associated cancers | Potential shedding pathways | Detection approaches in wastewater | References |
| --- | --- | --- | --- | --- | --- |
| Epstein–Barr virus (EBV) | dsDNA | Nasopharyngeal carcinoma; lymphomas | Saliva, faeces | qPCR; metagenomics; hybrid capture | Ahmed et al., 2023; Pongpakdeesakul, 2023; Peng et al., 2024; Prakash et al., 2026 |
| Hepatitis B virus (HBV) | dsDNA | Hepatocellular carcinoma | Blood, urine, faeces | qPCR; metagenomics | Hou et al., 2020; Peng et al., 2024 |
| Hepatitis C virus (HCV) | ssRNA | Hepatocellular carcinoma | Blood | Metagenomics | Stockdale et al., 2023 |
| Human herpesvirus 8 (HHV-8 / KSHV) | dsDNA | Kaposi sarcoma; primary effusion lymphoma; multicentric Castleman disease | Saliva, faeces, semen | qPCR; metagenomics | Miyani et al., 2021; Peng et al., 2024 |
| Human papillomavirus (HPV-16, HPV-18) | dsDNA | Cervical; anal; oropharyngeal; penile; vulvar cancers | Faeces, urine, genital secretions, skin | Multiplex PCR; sequencing; bead-based assays | La Rosa et al., 2013; Di Bonito et al., 2017 |
| Human T-lymphotropic virus type 1 (HTLV-1) | ssRNA | Adult T-cell leukemia/lymphoma | Blood, breast milk, semen | Hybrid capture | Prakash et al., 2026 |
| Merkel cell polyomavirus (MCPyV) | dsDNA | Merkel cell carcinoma | Skin, urine, faeces | Nested PCR; qPCR | Boffil-Mas et al., 2010; Di Bonito et al., 2015; Torres et al., 2016 |

In addition, MCPyV is a well-established etiological agent of the rare but aggressive Merkel cell carcinoma (Feng et al., 2008) (Table 1). Despite differences in transmission routes and disease manifestations, these viruses share several biological characteristics, including prolonged persistence in infected individuals, asymptomatic or latent infections, shedding of viral nucleic acids, and the potential for long-term circulation within human populations, all of which have important implications for population-level surveillance.

A defining characteristic of many oncogenic viruses is their ability to establish chronic or latent infections, often persisting for years or decades prior to the onset of malignancy (Butel, 2000). For example, EBV establishes lifelong latency, with malignancies arising under conditions of immune dysregulation (Young and Rickinson, 2004). These prolonged asymptomatic infections complicate conventional surveillance because infected individuals often remain undiagnosed and do not seek medical care, resulting in substantial underestimation of infection prevalence and delayed recognition of changes in viral circulation (de Martel et al., 2020). Such prolonged asymptomatic infections present challenges for conventional surveillance approaches (de Martel et al., 2020). Accordingly, the objective of wastewater surveillance is not to identify individuals at risk of cancer, but to monitor population-level circulation of oncogenic viruses independently of clinical testing and healthcare-seeking behaviour. However, there are opportunities to detect these viruses in municipal wastewater as viral fragments (nucleic acids) may be intermittently shed in faeces, urine, saliva, skin cells, genital secretions, or blood-derived materials during these prolonged infections. Viral particles, cell-associated DNA, and fragmented nucleic acids can enter sewage networks through a variety of human waste streams, generating aggregate population-level signals within wastewater systems (Prakash et al., 2026). Wastewater surveillance (WS) has emerged as a powerful, non-invasive approach for monitoring infectious diseases at the community scale, enabling the detection of pathogens independently of healthcare-seeking behaviour or access to clinical testing (Parkins et al., 2024). Although its public health applications differ between acute and chronic infections, the same underlying principle of monitoring community-level pathogen circulation is applicable to persistent oncogenic viruses.

The utility of WS was demonstrated extensively during the COVID-19 pandemic, when monitoring of SARS-CoV-2 RNA in wastewater provided early warning of community transmission and complemented clinical surveillance systems (Medema et al., 2020; Abunijela et al., 2026; Keller et al., 2026). Although the public health objectives differ between acute outbreak detection and surveillance of persistent oncogenic viruses, the COVID-19 pandemic established wastewater surveillance as a robust platform for monitoring pathogen circulation at the population level. Beyond respiratory pathogens, WS has been successfully applied to monitor enteric viruses, antimicrobial resistance markers, and other public health indicators. More recently, studies have reported the detection of oncogenic viruses, including HPV and EBV, in wastewater from Uruguay (Fernandez-Sabatella et al., 2025), Egypt (Hamza and Hamza, 2018), the United States (Prakash et al., 2026), Italy (Di Bonito et al., 2017), and Canada (Giesbrecht et al., 2025), demonstrating the feasibility of monitoring these viral targets at the community level. These findings highlight the potential of WS as a complementary tool for tracking the circulation of oncogenic viruses within populations, generating baseline data, identifying temporal and geographic trends in viral circulation, and evaluating the population-level impact of interventions such as vaccinations or screening programs, rather than serving as an early warning system for cancer incidence (Healy et al., 2026; Wade et al., 2022).

Wastewater surveillance of oncogenic viruses presents analytical challenges, including low target concentrations, intermittent shedding, and the presence of PCR inhibitors in complex wastewater matrices (Kitajima et al., 2020; Zafeiriadou et al., 2024). Nevertheless, molecular methods such as quantitative PCR and digital PCR enable sensitive detection and quantification of viral nucleic acids in environmental samples and are increasingly applied in WS studies (Kitajima et al., 2020; Clerkin et al., 2026). Despite these technological advances, WS efforts in Australia have largely focused on respiratory pathogens such as SARS-CoV-2, influenza viruses, and poliovirus, with limited attention given to oncogenic viruses. Consequently, there is little information on the presence, abundance, and temporal variability of these viruses in Australian wastewater systems. Establishing baseline data is important for assessing the feasibility of incorporating oncogenic virus targets into WS effortsand for supporting future population-level monitoring of viral circulation in Australia.

In the Australian context, the prevalence, cancer burden and mitigation strategies for these seven oncogenic viruses vary considerably, as summarized in Table 2. While established national programs have addressed HBV, HCV and HPV, surveillance and prevention for EBV, HHV-8, HTLV-1 and MCPyV remain limited. This variation underscores the potential value of community-level monitoring approaches, such as WS, to track the circulation of these viruses independently of clinical testing.

**Table 2.** Prevalence, virus-associated cancer incidence, and mitigation strategies for seven IARC Group 1 oncogenic viruses in Australia.

| Oncogenic viruses | Infection prevalence<sup>a</sup> | Incidence of Associated Cancer per 100,000<sup>b</sup> | Australian mitigation strategies<sup>c</sup> | References (a,b,c) |
| --- | --- | --- | --- | --- |
| Epstein–Barr virus (EBV) | 90% by adulthood | 3.2 Hodgkin lymphoma, NHL, NPC, Gastric cancer | No vaccine available. Research and surveillance focus on EBV-MS link via Ausimmune Study | <sup>a</sup>IARC, 1997<br><sup>b</sup>AIHW, 2023<br><sup>c</sup>Palmer et al., 2023; Lucas et al., 2022 |
| Hepatitis B Virus (HBV) | Australia ~0.8%<br>Queensland: ~0.4% a 0.7%,<br>Brisbane: ~0.6% a 0.9% | 10.0 Hepatocellular carcinoma | Universal infant + adolescent vaccination since 2000. ~85% coverage 2024 | <sup>a</sup>MacLachlan et al., 2025; ASHM, 2025<br><sup>b</sup>Canceraustralia, 2024<br><sup>c</sup>Department of Health, 2025 |
| Hepatitis C Virus (HCV) | Australia: ~0.27%<br>Queensland: ~0.25% | 4.0 Hepatocellular carcinoma | PBS-funded DAAs since 2016. National elimination strategy target 2030 | <sup>a</sup>ASHM, 2025<br><sup>b</sup>Howell, et al., 2025<br><sup>c</sup>Kwon et al 2019; Department of Health, 2025 |
| Human Herpesvirus (HHV-8) | Australia: Blood donors - 5.83% | 0.1 Kaposi Sarcoma | No vaccine. Indirect control via HIV management and targeted health initiatives for high-risk groups: MSM and Indigenous populations | <sup>a</sup>Speicher et al., 2022<br><sup>b</sup>IARC, 2022<br><sup>c</sup>Kirby Institute, 2023; AGDHAC, 2018; Healthdirect Australia - KS |
| Human papillomavirus (HPV) | Australia - HPV16/18 cervical cancer: 77%<br>QLD/Brisbane OPC 2019-2021: 68% | 6.7 Cervical cancer | National HPV vaccination program since 2007. Coverage 2024: 79.5% | <sup>a</sup>Yaksich et al., 2026<br><sup>b</sup>AIHW (2024)<br><sup>c</sup>Department of Health, 2025 |
| Human T-lymphotropic virus type 1 (HTLV-1) | Queensland - 0.1% Remote Indigenous communities of Central Australia > 30% Brisbane, seroprevalence - ~1% in the general population | 0.025 Adult T-cell leukemia/lymphoma | Blood safety approach: new-donor screening + universal leukoreduction instead of universal testing. Notifiable in NT and WA | <sup>a</sup>Noori et al., 2026; Einsiedel et al., 2021.<br><sup>b</sup>Martin et al., 2023<br><sup>c</sup>NT Health; Styles et al., 2017 |
| Merkel cell Polyomavirus (MCPyV) | Australia: 60% to 80% of adults | 1.6 Merkel Cell Carcinoma | No vaccine. Skin cancer prevention strategy: SunSmart programs + government shade initiatives to reduce UV exposure | <sup>a</sup>Carter et al., 2009; Viscidi et al., 2011<br><sup>b</sup>Youlden., 2024<br><sup>c</sup>Wong et al., 2015 |

**Abbreviations:** EBV, Epstein-Barr virus; HBV, Hepatitis B virus; HCV, Hepatitis C virus; HHV-8, Human herpesvirus 8; HPV, Human papillomavirus; HTLV-1, Human T-cell lymphotropic virus type 1; MCPyV, Merkel cell polyomavirus; PBS, Pharmaceutical Benefits Scheme; MSM, men who have sex with men; DAAs, direct-acting antivirals; NT, Northern Territory; QLD, Queensland; OPC, oropharyngeal cancer; WA, Western Australia; MS, multiple sclerosis; ~, approximate.
<sup>a</sup>: Infection prevalence estimates from national surveillance and seroprevalence studies. <sup>b</sup>: Age-standardized incidence rate per 100,000 persons of cancers attributable to the virus. <sup>c</sup>: Key national mitigation and surveillance strategies.

In this study, we applied PCR-based assays to detect and quantify seven IARC-classified oncogenic viruses in influent samples collected from six wastewater treatment plants across Southeast Queensland, Australia. This study provides baseline data on the occurrence and variability of oncogenic viruses in Australian wastewater and contributes to ongoing efforts to evaluate the potential for incorporating these targets into WS frameworks.

## 2. Materials and methods

### 2.1 Wastewater sampling

In total, seventy-six 24-h composite ∼1-L influent wastewater samples were collected from six municipal wastewater treatment plants (WWTPs; designated WWTP-A–F) located in Southeast, Queensland, Australia, between 31/12/2024 and 24/04/2025. This four-month sampling period was designed to provide a baseline assessment rather than capture seasonal variation in oncogenic virus occurrence. The selected WWTPs represent a range of municipal wastewater catchments and employ conventional activated sludge-based secondary treatment processes. To maintain confidentiality, all WWTPs were deliberately de-identified, and comparisons among individual catchments or treatment facilities were not intended. Influent samples were collected as flow-proportional 24-h composites using the routine sampling infrastructure at each facility. Samples were transferred to sterile containers, transported to the laboratory on ice, and initially processed as part of an ongoing WS program targeting other viral pathogens. Following primary analyses, aliquots of the raw influent wastewater were archived at −20°C. Archived samples were subsequently retrieved and analyzed in the present study.

### 2.2 Wastewater sample concentration

Archived raw influent wastewater samples stored at −20°C were thawed overnight at 4°C prior to viral concentration. An adsorption–extraction (AE) concentration method was subsequently used to concentrate viruses from the wastewater samples (Akter et al., 2024). In the AE workflow, MgCl_2_ (Sigma-Aldrich, St. Louis, Missouri, USA) was added to 50 mL aliquots of each wastewater sample to achieve a final concentration of 25 mM MgCl _2_. Each sample was then filtered through a 0.45 μm pore-size, negatively charged MCE membrane (47-mm diameter, HAWP04700, Darmstadt, Germany) using a magnetic filter funnel (Pall Corporation, Port Washington, New York, USA) and a filter flask (Merck Millipore Ltd.). After filtration, the membrane was aseptically removed from the filter funnel using sterilised tweezers, rolled, and inserted into a 7-mL bead-beating tube (Axygen, CA, USA). Each bead-beating tube contained approximately 1 g of zirconium oxide beads (0.5 mm and 1 mm diameter; 1:1 w/w; Next Advance, Inc., Troy, NY, USA) for nucleic acid extraction.

### 2.3 Nucleic acid extraction

Nucleic acid extraction from each influent composite wastewater sample was performed using the RNeasy PowerWater Kit (Cat. No. 14700–50-NF, Qiagen). To each bead-beating tube containing the membrane, 990 μL of lysis buffer PM1 (Qiagen, Hilden, Germany) and 10 μL of β-mercaptoethanol (Cat. No. M6250-10ML, Sigma-Aldrich, St. Louis, MO, USA) were added. The tube contents were homogenized using a Precellys 24 tissue homogenizer (Bertin Technologies, Montigny-le-Bretonneux, France) at 10,000 rpm for three 15 s cycles, with a 10 s interval between cycles. After homogenization, the tubes were centrifuged at 4,000 g for 5 min to pellet filter debris, solids, and beads. DNase I solution was omitted from the protocol to enable simultaneous isolation of both DNA and RNA targets, supporting multi-pathogen detection. The sample was then eluted with 150 μL of nuclease-free water and stored at −20°C prior to qPCR/RT-PCR analysis. Concentrations and quality (*A*_260/280_ and *A*_260/230_) of nucleic acid samples were measured using a DeNovix DS-11 Series Spectrophotometer/Fluorometer (Wilmington, DE, USA).

### 2.4 PCR inhibition testing

A Sketa22 PCR assay was employed to determine PCR inhibition by seeding a known copy number (10^4^) of *Oncorhynchus keta (O. keta)* DNA in extracted wastewater nucleic acid samples (Haugland et al., 2005). The reference quantification cycle (Cq) value was determined from PCR reaction containing only the positive control and compared with the Cq values obtained from all nucleic acid samples.

Samples were considered to have no PCR inhibition when the Cq values of nucleic acid samples were within 2 Cq values of the reference Cq value (Staley et al., 2012). A DNA-based inhibition control was selected because the majority of the oncogenic viral targets investigated in this study (5 of 7) were DNA viruses.

### 2.5 PCR/RT-PCR assays

The detection HPV-16 E7 gene (Bordigoni et al., 2021) and HPV-18 E7 gene (Bordigoni et al., 2021) was performed using published PCR assays (Supplementary Table ST1). The PCR (i.e., real-time) analyses were conducted in 20 μL reactions mixtures containing 10 μL of 2x QuantiNova Probe PCR Master Mix (Qiagen, Hilden Germany), 800 nM of each forward and reverse primer, 200 nM of probe, and 5 μL of nucleic acid extract. The detection of HCV and HTLV-1 were performed using RT-PCR assays in 20 μL reaction mixtures containing 5 μL of TaqMan™ Fast Virus 1-Step Master Mix (Applied Biosystem; California, USA), 800 nM of each forward and reverse primer, 200 nM of probe, and 5 μL of nucleic acid extract. Target, gene, primer and probe sequences, assay types and thermal cycling parameters are detailed in Supplementary Table ST1. Synthetic gBlocks were used as positive controls. All PCR and RT-PCR reactions were performed in triplicate on a Bio-Rad CFX Opus Real-time PCR system (Bio-Rad Laboratories, Hercules, California, USA). Each run included positive controls and a no-template control (NTC), and threshold and baseline settings were determined automatically by the instrument software.

### 2.6 dPCR assays

The quantification HBV (Wenzel et al., 2006), EBV (Kimura et al., 1999), HHV-8 (Miyani et al., 2021), HPV-16 E6 (Bordigoni et al., 2021), HPV-18 E6 (Bordigoni et al., 2021), and MCPyV (Sadeghi et al., 2012) was performed using published PCR assays (Supplementary Table ST1). dPCR analyses were conducted using the QIAcuity Probe PCR Kit (Cat No. 250103; Qiagen) and 26K 24-well Nanoplates (Cat No. 250001; Qiagen). The QIAcuity 26K 24-well Nanoplates generate 26,000 partitions per well (0.91 nL/partition). dPCR reactions (40 µL) contained 10 µL of QIAcuity Probe PCR Master Mix, 800 nM each of the forward and reverse primers, 400 nM probe, and 5-10 µL of template nucleic acid. One dPCR replicate was analyzed for each sample.

The 40-µL dPCR reactions were prepared in a 96-well pre-plate and then transferred to the 26K 24-well Nanoplate. Nanoplates were loaded onto the QIAcuity dPCR 5-plex platform (Qiagen) for partition generation, thermal cycling and fluorescence imaging in the FAM and HEX channels. Each dPCR run included a single positive control (gBlocks) and NTC. Data were analysed using QIAcuity Software Suite v3.2.0.0, and target concentrations were exported as gene copies (GC)/µL of template. Thresholds were determined using the software’s automatic settings. A combination of qPCR and dPCR platforms was used to optimise resource efficiency and analytical throughput across targets.

### 2.7 QA/QC and data interpretation

To minimize contamination, nucleic acid extraction and PCR setup were performed in physically separate laboratories. Runs were considered valid only when no amplification was observed in the no-template controls and all positive controls performed as expected. To assess the integrity of nucleic acids following storage at −20°C, *Carjivirus* was quantified in all wastewater samples as an endogenous viral process control. As *Carjivirus* is highly abundant and consistently present in human wastewater, measured concentrations were used to verify that substantial nucleic acid degradation had not occurred during sample storage prior to analysis. For PCR and RT-PCR assays (i.e., qualitative detection), wastewater samples were classified as positive when amplification was detected in at least one of three technical replicates within 40 cycles. Samples with Cq values between 40 and 45 were classified as trace detections. Samples were considered not detected (ND) when no amplification was observed in any replicate. For dPCR assays, samples were classified as positive when ≥1 positive partition was detected, and no positive partitions were observed in the corresponding no-template control. Samples were considered quantifiable when ≥3 positive partitions were observed. dPCR runs were considered valid when >20,000 accepted partitions were generated per sample.

## 3. Results

### 3.1 QA/QC and PCR inhibition

No amplification was observed in the no-template controls across all assays. All positive controls resulted in positive detection, as expected. The number of accepted partitions exceeded 20,000 for all reactions, providing sufficient resolution for reliable target detection and quantification. Clear separation of positive and negative partitions was observed across all assays. PCR inhibition testing using the Sketa22 assay indicated no evidence of PCR inhibition in any samples, with al l Cq values within ±2 cycles of the reference control. The mean *Carjivirus* concentration in all samples was 8.49 ± 0.79 log₁₀ GC/L, which is within the range commonly reported for municipal wastewater. These findings suggest that sample integrity was maintained during storage.

### 3.2 Detection frequency of oncogenic viruses in influent wastewater

A total of 76 influent wastewater samples collected across six WWTPs were screened for oncogenic viruses (Fig. 1 and Supplementary Table ST2). Detection frequencies varied substantially among viral targets. MCPyV was the most frequently detected virus, identified in 97.3% (74/76) of samples. Detection of MCPyV was consistent across al l WWTPs, with 100% positivity observed at WWTP-A, WWTP-B, WWTP-E, and WWTP-F, and slightly lower detection at WWTP-D (93.3%) and WWTP-C (91.7%). HBV was the second most frequently detected virus, identified in 26.3% (20/76) of samples. HBV detection frequencies varied across WWTPs, ranging from 13.3% at WWTP-D to 54.5% at WWTP-E. EBV was detected in 15.8% (12/76) of samples, with site-specific detection frequencies ranging from 9.1% to 30.8%. HPV-16 and HPV-18 were detected at lower frequencies. HPV-16 and HPV-18 targets were detected in 9.2% (7/76) and 10.5% (8/76) of samples, respectively. Detection of HPV-16 and HPV-18 was uneven across WWTPs, with most positive detections occurring at WWTP-A and WWTP-B and no detections at WWTP-D. HHV-8 was detected in 3.9% (3/76) of samples, with positive samples detected at WWTP-A and WWTP-C. HTLV-1 and HCV were each detected in a single sample (1.3%), at WWTP-C and WWTP-D, respectively.

**Fig. 1.**
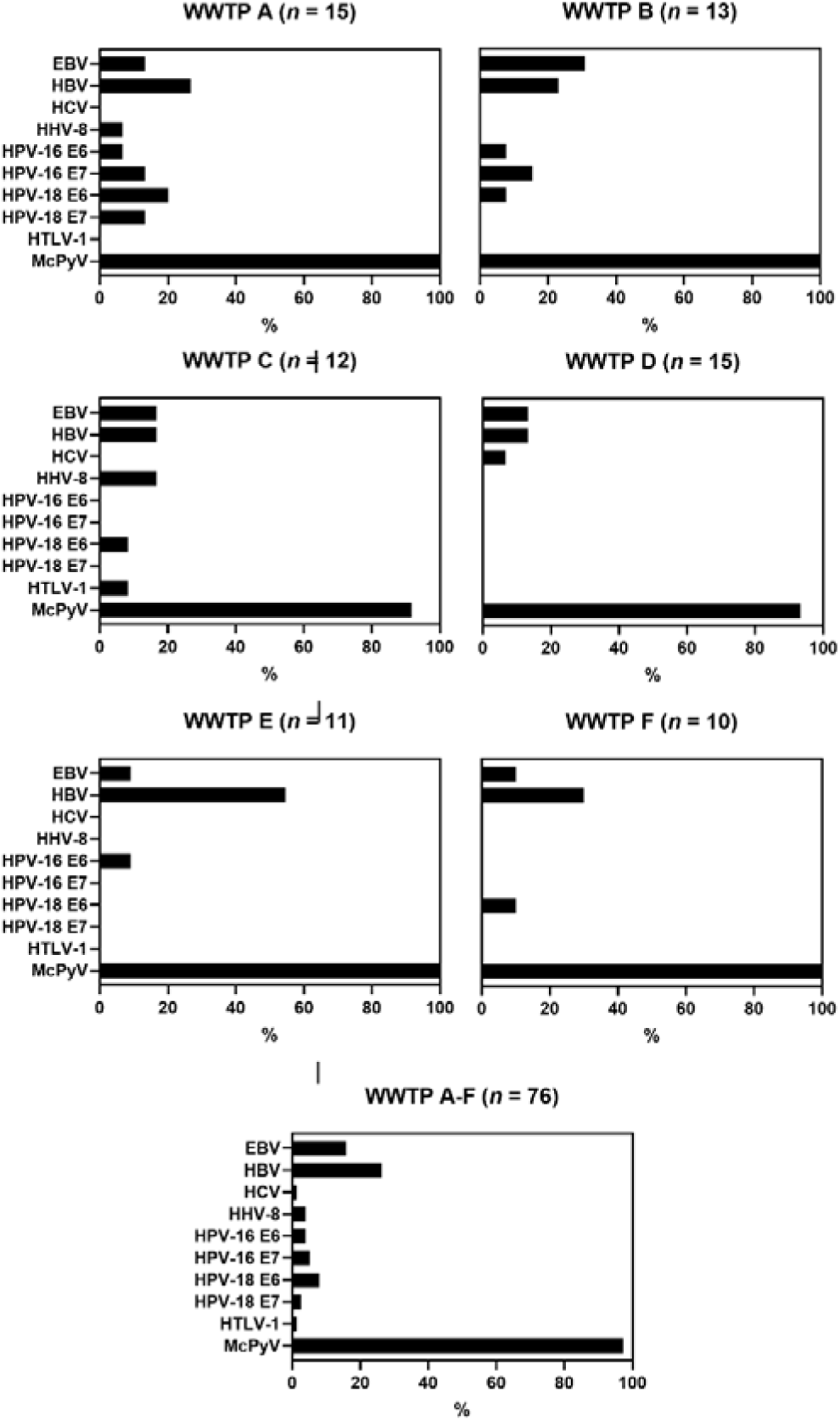
Detection rate (%) of 10 oncogenic viruses in the influent of 6 Wastewater Treatment Plants - WWTPs in Southeast Queensland, Australia. Bar graphs show the percentage of positive samples for EBV, HBV, HCV, HHV-8, HPV-16 E6/E7, HPV-18 E6/E7, HTLV-1 and MCPyV in each WWTP. The bottom panel presents consolidated data from all 6 WWTPs. n = total number of samples per WWTP.

### 3.3 Concentrations of oncogenic viruses in influent wastewater

Quantifiable concentrations of oncogenic viral targets were obtained using digital PCR (dPCR) in a subset of influent wastewater samples (Table 3). Overall, MCPyV was the most frequently quantified viral target and was detected above the limit of quantification at all six wastewater treatment plants (WWTPs). At WWTP-A, MCPyV was quantified in all samples (15/15; 100%) with a mean concentration of 3.52 ± 0.46 log₁₀ GC/50 mL. HBV and HPV-16 E6 were each quantified in a single sample (1/15; 6.66%), with concentrations of 2.00 log₁₀ GC/50 mL and 2.64 log₁₀ GC/50 mL; all remaining samples were either not detected or below the quantification threshold. At WWTP-B, MCPyV was quantified in 12 of 13 samples (92.3%), with a mean concentration of 3.23 ± 0.59 log₁₀ GC/50 mL. HBV was quantified in one sample (1/13; 7.69%) at 2.03 log₁₀ GC/50 mL.

**Table 3.** Quantifiable concentrations of oncogenic viral targets in influent wastewater as determined using Dpcr.

| WWTPs | Target | No. of quantifiable samples/No. of samples tested (%) | Mean log<sub>10</sub> GC/50 mL ± SD |
| --- | --- | --- | --- |
| WWTP-A | HBV | 1/15 (6.66) | 2.00 |
|  | HPV 16-E6 | 1/15 (6.66) | 2.64 |
|  | MCPyV | 15/15 (100) | 3.52 ± 0.46 |
| WWTP-B | HBV | 1/13 (7.69) | 2.03 |
|  | MCPyV | 12/13 (92.3) | 3.23 ± 0.59 |
| WWTP-C | HBV | 1/12 (9.09) | 1.95 |
|  | MCPyV | 11/12 (91.7) | 3.85 ± 0.48 |
| WWTP-D | EBV | 1/15 (6.66) | 2.58 |
|  | MCPyV | 13/15 (93.3) | 3.09 ± 0.36 |
| WWTP-E | EBV | 1/11 (9.09) | 2.90 |
|  | HBV | 1/11 (9.09) | 2.11 |
|  | MCPyV | 11/11 (100) | 3.35 ± 0.49 |
| WWTP-F | MCPyV | 9/10 (90) | 3.32 ± 0.59 |

At WWTP-C, MCPyV was quantified in 11 of 12 samples (91.7%), with a mean concentration of 3.85 ± 0.48 log₁₀ GC/50 mL. HBV was quantified in a single sample (1/12; 9.09%) at 1.95 log₁₀ GC/50 mL. At WWTP-D, MCPyV was quantified in 13 of 15 samples (93.3%), with a mean concentration of 3.09 ± 0.36 log₁₀ GC/50 mL. EBV was quantified in one sample (1/15; 6.66%) at 2.58 log₁₀ GC/50 mL. At WWTP-E, MCPyV was quantified in all samples (11/11; 100%), with a mean concentration of 3.35 ± 0.49 log₁₀ GC/50 mL. EBV and HBV were each quantified in one sample (1/11; 9.09%), with concentrations of 2.90 log₁₀ GC/50 mL and 2.11 log₁₀ GC/50 mL, respectively. At WWTP-F, MCPyV was quantified in 9 of 10 samples (90%), with a mean concentration of 3.32 ± 0.59 log₁₀ GC/50 mL. No other viral targets were quantified above the limit of quantification at this site. Overall, MCPyV was consistently quantifiable across all wastewater treatment plants, whereas HBV, EBV, and HPV-16 were quantifiable only sporadically and in a limited number of samples.

### 3.4 Spatial variability and co-occurrence among WWTPs

Detection patterns varied among WWTPs, indicating spatial variability in the occurrence of oncogenic viruses (Table 3 and Supplementary Table ST2). WWTP-A exhibited the highest viral diversity of detected viral targets, with seven viral targets detected at least once during the study period. In comparison, six targets were detected at WWTP-B, WWTP-C, four at WWTP-D, WWTP-E, and WWTP-F (Fig 1 and Table ST3). MCPyV was detected at all WWTPs and was the dominant viral target in terms of both detection frequency and quantifiable occurrence. HBV and EBV were detected across multiple WWTPs, although at generally lower frequencies compared with MCPyV. HPV-16 and HPV-18 showed a more restricted distribution and were detected primarily at WWTP-A and WWTP-B, with limited detections at other sites. HHV-8, HTLV-1, and HCV were detected sporadically and were confined to one or two WWTPs. Multiple viral targets were detected within individual wastewater samples. MCPyV was frequently detected alongside other viral targets, including HBV and EBV, although formal co-occurrence analyses were not conducted. No consistent co-occurrence patterns were observed among the less frequently detected viruses. Overall, no apparent relationship was observed between WWTP population size and viral target diversity of detections.

## 4. Discussion

This study represents the first comprehensive assessment of oncogenic viral markers in municipal wastewater in Australia and demonstrates the potential of WS to detect and monitor viruses associated with infection-related cancers at the population level. Seven IARC-classified oncogenic viral targets were screened in influent wastewater collected from six wastewater treatment plants (WWTPs) serving the Southeast Queensland region. Viral nucleic acids from all seven targets were detected at least once during the study period, although detection frequencies varied considerably among viruses. These differences may be influenced by several factors, including differences in population prevalence, viral shedding pathways and rates, viral loads released into wastewater, environmental persistence, and analytical sensitivity of the assay used.

Prakash et al. (2026) similarly reported the detection of multiple oncogenic viruses in wastewater across 16 cities in Texas during a three-year surveillance program, demonstrating the feasibility of monitoring oncogenic viral markers at large geographic scales. Consistent with their findings, several oncogenic viruses were detected across multiple WWTPs in the present study despite differences in catchment population size and characteristics. The detection of oncogenic viral markers across geographically distinct populations in both Australia and the United States indicates that municipal wastewater contains measurable signals from a diverse range of oncogenic viruses. These findings support the feasibility of incorporating selected oncogenic viral targets into WS programs to establish baseline patterns of viral circulation, monitor long-term temporal trends, and evaluate the population-level impact of public health interventions where applicable, such as HPV and HBV vaccination programs. For viruses without effective preventive interventions, such surveillance may provide baseline epidemiological data and improve understanding of population-level viral circulation.

Among the viral targets investigated, MCPyV was the most consistently detected and quantifiable marker, occurring in 97.3% of wastewater samples and quantified in the majority (93.4%) of samples from all six WWTPs. This widespread occurrence is consistent with the high global seroprevalence of MCPyV and its near-ubiquitous distribution in human populations. MCPyV is recognised as the principal aetiological agent of Merkel cell carcinoma (MCC), a rare but aggressive skin cancer, although, infection is common and typically asymptomatic in immunocompetent individuals (Kean et al., 2009; Tolstov et al., 2009). While the public health utility of routine wastewater surveillance for MCPyV remains uncertain given the rarity of MCC, our findings establish baseline occurrence and concentration data for this ubiquitous MCPyV. Such baseline data may prove valuable for understanding environmental shedding patterns and for informing future assessments of whether MCPyV has utility as a biomarker in specific epidemiological or immunocompromised populations.

The high detection frequency of MCPyV observed in this study is consistent with previous wastewater investigations. For example, Di Bonito et al. (2015) detected MCPyV in 50.3% of untreated wastewater samples from WWTPs in Italy. Differences in detection frequencies between studies may reflect variations in analytical sensitivity, sample processing methods, wastewater characteristics, and temporal patterns of viral shedding (Hart and Halden, 2020; Kitajima et al., 2020). Although Queensland reports the highest incidence rates of MCC globally (Youlden et al., 2014), the relationship between wastewater MCPyV concentrations and MCC incidence remains unclear. MCPyV infection is widespread in the general population, whereas MCC is a rare cancer that develops only in a small subset of infected individuals. Therefore, MCPyV concentrations in wastewater likely reflect overall viral circulation and shedding rather than disease incidence. Further studies linking wastewater MCPyV measurements with epidemiological data would be required to assess whether any population-level association exists.

HBV and EBV were detected at moderate frequencies across multiple WWTPs, although quantifiable concentrations were obtained only sporadically. In the Australian context, HBV has a low prevalence of approximately 0.8% nationally and 0.4–0.7% in Queensland, with a cancer incidence of 10.0 per 100,000 (Cancer Australia, 2024). Mitigation includes universal infant vaccination since 2000 and ∼85% coverage in 2024. EBV has very high prevalence of ∼90% by adulthood and a cancer incidence of 3.2 per 100,000, with no licensed vaccine and ongoing research via the Ausimmune Study.

HBV and EBV were detected across all six WWTPs, with HBV detected more frequently than EBV (26.3% vs. 15.8%), although quantifiable concentrations of both viruses were obtained only sporadically. Consistent with our findings, prior studies have reported low HBV detection in wastewater. Farkas et al. (2022) did not detect HBV in 115 samples from 13 UK WWTPs, attributing this to environmental degradation. Peng et al. (2024) detected HBV in 1/12 samples using metagenomics and qPCR, despite high prevalence of HBV in China, likely reflecting limited sensitivity of qPCR assay for detecting low-abundance viral targets. In contrast, Li et al. (2024) reported *Orthohepadnavirus* in 19% (9/48) of samples in Detroit, Michigan, while Prakash et al. (2026) observed low-level detection across 16 cities in Texas. Collectively, these studies, along with our findings, suggest that HBV occurs in wastewater at low frequency and abundance relative to the other target human viruses. However, it remains unclear whether this primarily reflects low viral concentrations resulting from limited shedding into wastewater, rapid environmental degradation, or methodological limitations in detecting low-abundance viral targets.

EBV has been previously detected in wastewater in Australia (Ahmed et al., 2023) and the United States (Prakash et al., 2026). Because EBV is shed in saliva and oropharyngeal secretions, continuous viral input into wastewater could occur through daily activities of infected individuals. The high global prevalence of EBV (Hoover and Higginbotham, 2006), together with its lifelong persistence and episodic reactivation, likely explains its consistent detection in wastewater despite the relatively low concentrations observed. However, the relatively low concentrations observed may reflect the intermittent and highly variable nature of EBV shedding, the predominance of shedding via saliva rather than faeces, and the dilution of viral material within wastewater systems. In addition, because wastewater concentrations were generally low, analytical sensitivity may also have influenced measured concentrations and detection frequencies.

For this study, HPV-16 and HPV-18 were selected as they represent the most clinically significant high-risk oncogenic genotypes, with E6 and E7 oncoproteins mediating p53 and Rb inactivation and driving malignant transformation. Both genotypes were infrequently detected in influent wastewater, likely reflecting the biology of HPV infection and its transmission via direct mucosal contact rather than through feces. Furthermore, viral nucleic acid entering wastewater may be fragmented and present at very low concentrations, reducing analytical sensitivity. In Australia, HPV has moderate prevalence with an associated cancer incidence of 7.0 per 100,000. Mitigation includes a national school-based vaccination program with ∼80% coverage in females and ∼76% in males in 2022.

Previous studies have nevertheless reported widespread HPV detection in wastewater globally, including in Egypt (Hamza and Hamza, 2018; Ahmed et al., 2019), Uruguay (Victoria et al., 2022), and Europe (Di Bonito et al., 2017), with detection frequencies of 30–73% and identification of both high-and low-risk genotypes. The lower detection frequency observed in our study likely reflects methodological differences, as prior studies commonly applied broad-range PCR, genotyping, or sequencing approaches capturing multiple HPV types, whereas this study targeted only HPV-16 and HPV-18. Second, Australia’s high uptake of prophylactic HPV vaccination has substantially reduced the prevalence of vaccine-targeted HPV-16 and HPV-18 infections, which may also contribute to their lower detection in wastewater. Despite this narrower scope and low environmental abundance, detection of both targets in our study demonstrates the feasibility of tracking clinically relevant oncogenic HPV in wastewater, although improved sensitivity and broader multi-target strategies are required for robust environmental surveillance.

HHV-8, HTLV-1, and HCV were detected sporadically (<3.50% detection frequency across 76 samples) in the present study, with most positive samples below the limit of quantification threshold. This low detection aligns with their epidemiology in Australia: HCV prevalence is ∼0.27% with cancer incidence of 4.0 per 100,000 and mitigation via PBS-funded DAAs since 2016 and a 2030 elimination target. HHV-8 has an estimated prevalence of 5.83% in blood donors and cancer incidence of 0.1 per 100,000, with mitigation focused on HIV management. Nevertheless, their detection is consistent with previous wastewater studies reporting their presence in municipal wastewater. HCV has been identified in untreated wastewater using both metagenomic and targeted molecular approaches, although detection frequencies have varied considerably (11.1% - 80.7%) among studies (McCall et al., 2020; Stockdale et al., 2023; Alshehri et al., 2025). Similarly, HHV-8 has been reported in wastewater from Detroit, USA, where it was detected at high frequency (94.4%) and abundance using a combination of metagenomic sequencing and qPCR (Miyani et al., 2020). In contrast, Prakash et al. (2026) observed frequent non-detection or low read abundance for HCV, HHV-8 (KSHV), and HTLV-1 in wastewater samples collected in Texas. Given the low concentrations and detection frequencies observed, further work is needed to evaluate the utility of these viruses as WS targets and to clarify the relationship between wastewater signals and population-level infection burden.

Several considerations are important for interpreting the findings of this study. First, samples were analyzed from six WWTPs within a single metropolitan region, limiting the generalisability of our findings to broader geographic and demographic settings across Australia. Furthermore, our analysis measured viral nucleic acids in wastewater; therefore, the results reflect the presence and/or concentrations of viral genetic material rather than infectious virus, precluding any direct inferences regarding viral infectivity. Resolved, geocoded epidemiological data were not incorporated into this study, preventing assessment of associations between wastewater signals and infection prevalence in the contributing populations. Moreover, the sampling period covered approximately four months and therefore did not encompass a full annual cycle. Consequently, potential seasonal patterns in the circulation, shedding, and wastewater detection of some oncogenic viruses could not be assessed. Future longitudinal studies spanning multiple seasons or years are needed to determine whether temporal variability influences wastewater detection frequencies and concentrations.

Additionally, different molecular platforms (real-time PCR/RT-PCR and dPCR) were used for different viral targets. Although validated published assays were employed, differences in analytical sensitivity among assays and platforms may have influenced detection frequencies, particularly for low-abundance targets. Because assay-specific limits of detection (LoDs) were not determined under the conditions of this study, comparisons of detection frequencies among viral targets should be interpreted with caution. Finally, several viral targets were detected only rarely, limiting the ability to assess temporal patterns and spatial trends. Additional work is required to better characterise the behaviour of oncogenic viruses in wastewater systems, which will be critical for accurate interpretation of WS data. Future studies should also integrate wastewater surveillance with clinical and epidemiological data collected over extended monitoring periods to better understand the relationship between wastewater signals and population-level infection dynamics.

Overall, this study provides the first evidence that multiple oncogenic viral markers can be detected and quantified in Australian wastewater using PCR and dPCR. The consistent detection of MCPyV and the repeated detection of HBV, EBV, and HPV markers demonstrate that wastewater contains measurable signals from viruses associated with infection-related cancers. These findings establish a baseline for future research and can be used to prioritise oncogenic viral targets for wastewater surveillance and methodological development. As WS continues to evolve beyond surveillance of respiratory viruses, the integration of oncogenic viruses may offer new opportunities for understanding long-term patterns of viral circulation and inform broader public health strategies for cancer prevention.

## 4. Conclusions

- This study provides the first baseline assessment of oncogenic viruses in Australian wastewater, demonstrating that all targeted viruses (i.e., EBV, HBV, HCV, HHV-8, HPV-17, HPV-18, HTLV-1 and MCPyV) were detectable at least once across six WWTPs in Southeast Queensland.
- MCPyV was consistently detected across all WWTPs, appearing in 97.3% of samples, confirming its widespread circulation and strong suitability as a WS target.
- HBV and EBV were detected intermittently across multiple catchments, while HPV-16/18, HHV-8, HTLV-1, and HCV occurred at low frequencies, likely reflecting differences in population prevalence and/or shedding dynamics.
- The successful detection of both common (i.e., McPyV, HBV, amd EBV) and rare (i.e., HPV-16/18. HHV-8. HTLV-1, and HCV) oncogenic viruses highlights the sensitivity of WS for capturing community-level infection signals. These baseline data establish a foundation for future research evaluating the feasibility and value of incorporating oncogenic viral markers into broader public health and cancer-related surveillance efforts.

## Data Availability

All data produced in the present work are contained in the manuscript

---

**Supplementary Table ST.**
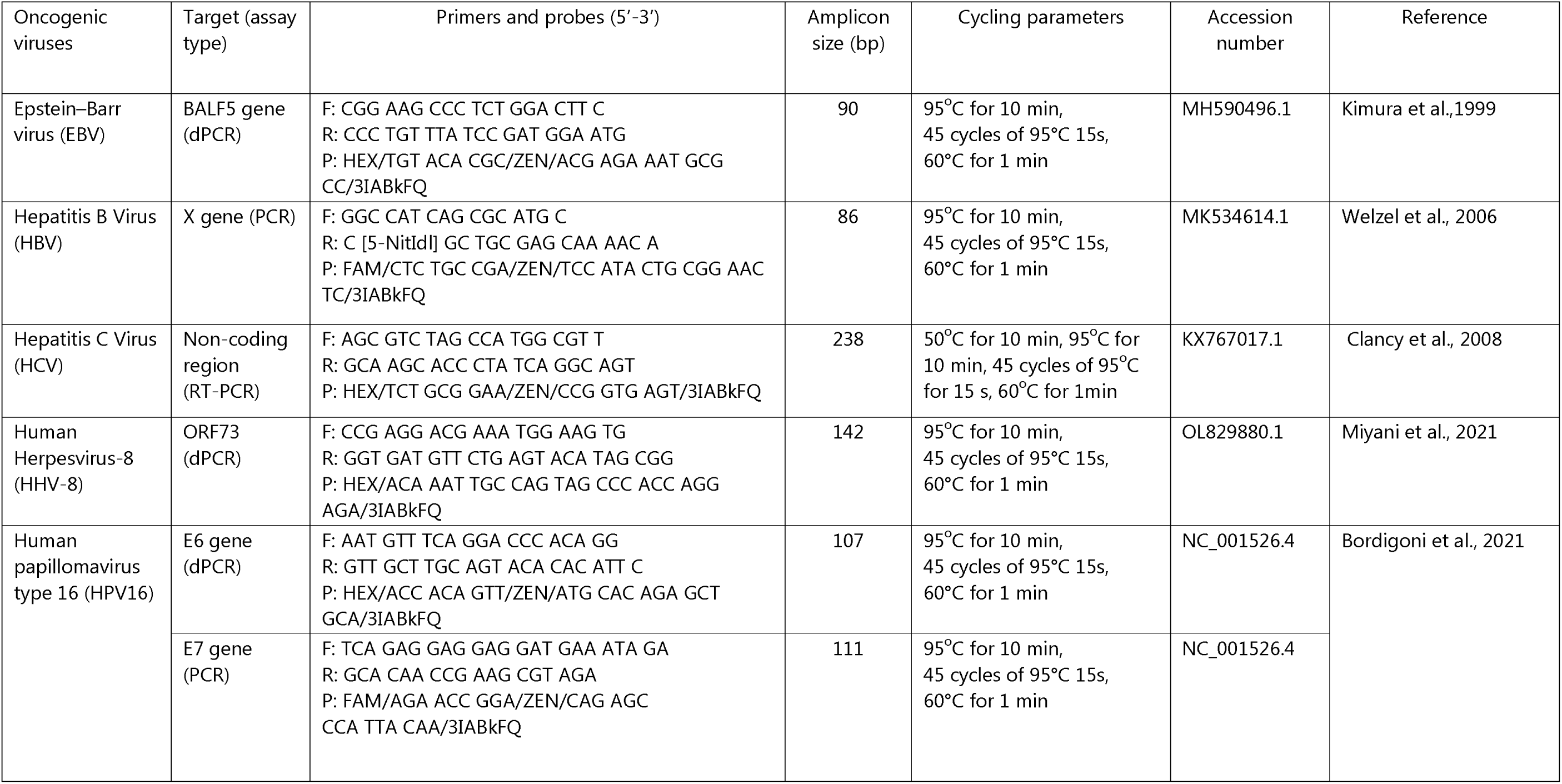
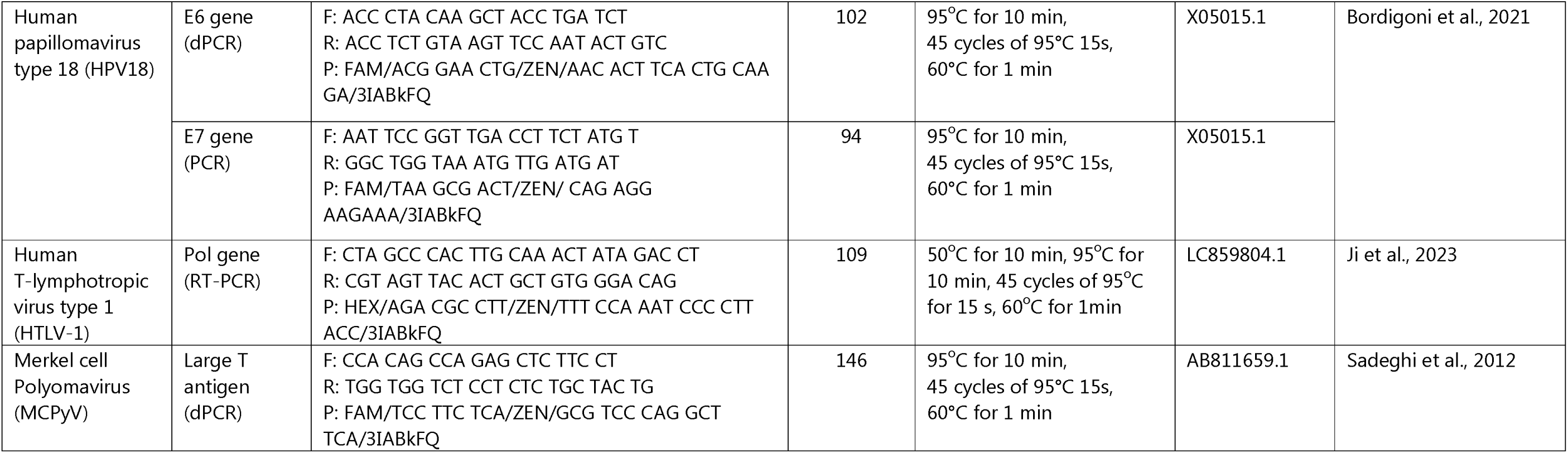
Primer and probe sequences and thermal cycling conditions.

**Supplementary Table ST2.** Detection frequency of oncogenic virus genomes in influent wastewater samples collected from six wastewater treatment plants (WWTPs) in Southeast Queensland, Australia.

| WWTP | No. of positive sample/total number of samples (% positive) |  |  |  |  |  |  |  |  |  |
| --- | --- | --- | --- | --- | --- | --- | --- | --- | --- | --- |
|  | EBV | HBV | HCV | HHV-8 | HPV-16 E6 | HPV-16 E7 | HPV-18 E6 | HPV-18 E7 | HTLV-1 | MCPyV |
| WWTP-A | 2/15 (13.3) | 4/15 (26.7) | 0/15 (0) | 1/15 (6.66) | 1/15 (6.66) | 2/15 (13.3) | 3/15 (20.0) | 2/15 (13.3) | 0/15 (0) | 15/15 (100) |
| WWTP-B | 4/13 (30.8) | 3/13 (23.1) | 0/13 (0) | 0/13 (0) | 1/13 (7.69) | 2/13 (15.4) | 1/13 (7.69) | 0/13 (0) | 0/13 (0) | 13/13 (100) |
| WWTP-C | 2/12 (16.7) | 2/12 (16.7) | 0/12 (0) | 2/12 (16.7) | 0/12 (0) | 0/12 (0) | 1/12 (8.33) | 0/12 (0) | 1/12 (8.33) | 11/12 (91.7) |
| WWTP-D | 2/15 (13.3) | 2/15 (13.3) | 1/15 (6.66) | 0/15 (0) | 0/15 (0) | 0/15 (0) | 0/15 (0) | 0/15 (0) | 0/15 (0) | 14/15 (93.3) |
| WWTP-E | 1/11 (9.09) | 6/11 (54.5) | 0/11 (0) | 0/11 (0) | 1/11 (9.09) | 0/11 (0) | 0/11 (0) | 0/11 (0) | 0/11 (0) | 11/11 (100) |
| WWTP-F | 1/10 (10.0) | 3/10 (30.0) | 0/10 (0) | 0/10 (0) | 0/10 (0) | 0/10 (0) | 1/10 (10.0) | 0/10 (0) | 0/10 (0) | 10/10 (100) |
| Total | 12/76 (15.8) | 20/76 (26.3) | 1/76 (1.31) | 3/76 (3.93) | 3/76 (3.93) | 4/76 (5.26) | 6/76 (7.89) | 2/76 (2.63) | 1.76 (1.31) | 74/76 (97.3) |

